# Aggressive Antipyretics in CNS Malaria: Study Protocol of a Randomized-Controlled Trial Assessing Antipyretic Efficacy and Parasite Clearance Effects (Malaria FEVER Study)

**DOI:** 10.1101/2022.05.05.22274544

**Authors:** Moses Chilombe, Michael McDermott, Karl B. Seydel, Manoj Mathews, Musaku Mwenechanya, Gretchen L. Birbeck

## Abstract

**Background:** Despite ongoing eradication efforts, malaria remains a major public health challenge in Africa where annually, ~250,000 children with malaria experience a neurologic injury with subsequent neurodisability. Evidence indicates that among children with CNS malaria, a higher temperature during the acute illness is a risk factor for post-infectious neurologic sequelae. As such, aggressive antipyretic therapy may be warranted, at least among children with complicated malaria who are at substantial risk of brain injury. Previous clinical trials conducted primarily in children with uncomplicated malaria and using only a single antipyretic medication have shown limited benefits in terms of fever reduction; however, no studies to date have examined malaria fever management using dual therapies. In this clinical trial of aggressive antipyretic therapy, children hospitalized with CNS malaria will be randomized to usual care (acetaminophen every 6 hours for a temperature ≥ 38.5°C) vs. prophylactic acetaminophen and ibuprofen every 6 hours for 72 hours. This proof-of-concept study will determine whether aggressive antipyretic therapy results in a lower mean maximum temperature relative to usual care.

**Methods:** We will enroll 284 participants from three settings at Queen Elizabeth Central Hospital in Blantyre, Malawi; at the University Teaching Hospitals Children’s Hospital in Lusaka, Zambia and at Chipata Central Hospital, Chipata, Zambia. Parents or guardians must provide written informed consent. Eligible participants are 2-11 years with evidence of P. falciparum malaria infection by peripheral blood smear or rapid diagnostic test with CNS symptoms associated with malaria. Eligible children will receive treatment allocation as determined by randomization and will be assigned to treatment groups with 1:1 allocation using blocked randomization.

**Discussion:** The clinical trial proposed here seeks to challenge the practice paradigm of limited fever treatment based upon hyperpyrexia by evaluating the fever-reduction efficacy of more aggressive antipyretic use involving two antipyretics and prophylactic administration while also taking advantage of a relatively new method for quantifying total parasite burden (HRP2 quantification) to further characterize malaria severity and elucidate the impact of antipyretics on parasite sequestration and clearance. If aggressive antipyretic therapy is shown to safely reduce the maximum temperature during CNS malaria, a clinical trial evaluating the neuroprotective effects of temperature reduction in CNS malaria is warranted.

**Trial registration:** This trial is registered with ClinicalTrial.gov (NCT03399318) and with the Pan African Clinical Trials Registry (PACTR201804003255157)

## Introduction

### Background and rationale

#### Description of research question

##### Significance

###### Neurologic Sequelae Following Severe Malaria in Childhood

Despite eradication measures, ~200 million malaria cases occurred in 2013 with ~500,000 malarial deaths, primarily in African children(1). Neurologic sequelae occur in 1/3^rd^ of survivors of CNS malaria, meaning parasitemia and neurologic symptoms including impaired consciousness and/or seizures(2–5). Newer antimalarial medications rapidly clear peripheral parasitemia and improve survival, but mortality remains high at 12-25% with no associated decline in post-malaria neurologic injury (6). Past studies have identified neurologic deficits at discharge in 9-18% of CNS malaria survivors with additional sequelae, such as epilepsy, becoming evident with longer term follow-up (2, 7–13). We conducted a prospective cohort study (BMPES) of children with the most severe form of CNS malaria—cerebral malaria (CM). CM is defined as unarousable coma (persisting for at least 30 minutes after any seizure), parasitemia and no other coma etiology(14). When followed for an average of 77 weeks, 32% of BMPES children experienced neurologic sequelae including epilepsy (9%), behavioral problems (11%) and new neurodisabilities (23%)(2). In Table 1 below, data from studies in other malaria-endemic African countries indicate that neurologic sequelae are not unique to children with CM. Children with “CNS malaria”, meaning malaria-associated seizures or impaired consciousness without deep coma also experience high rates of sequelae.

**Table 1:**
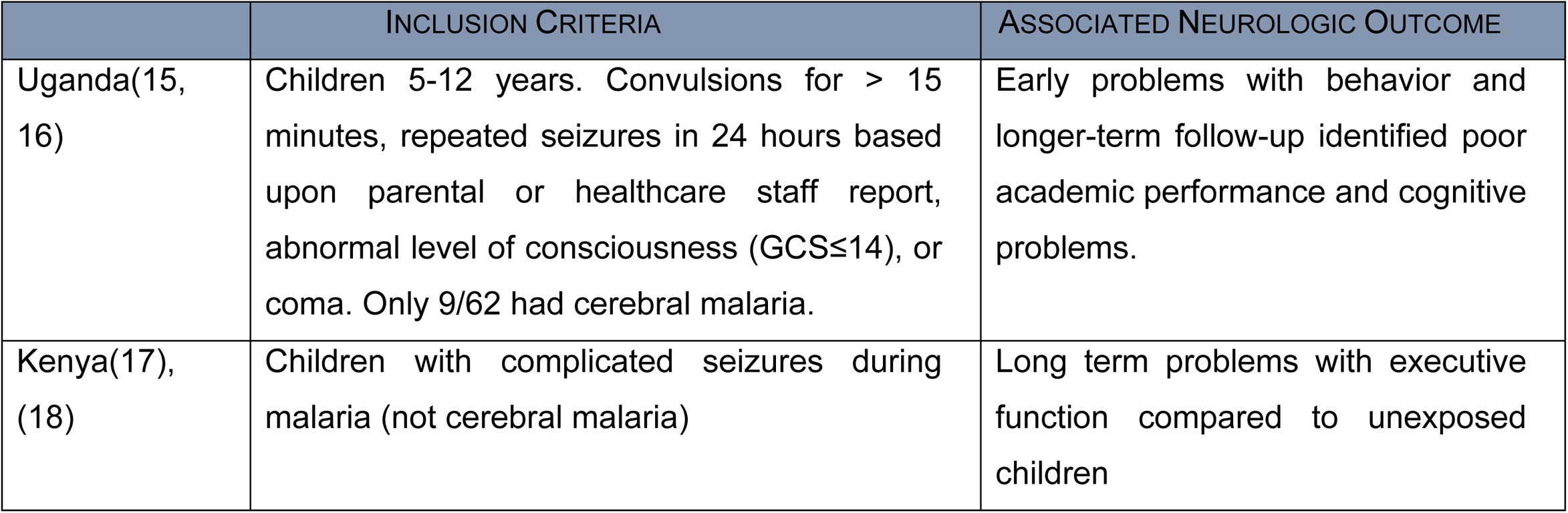
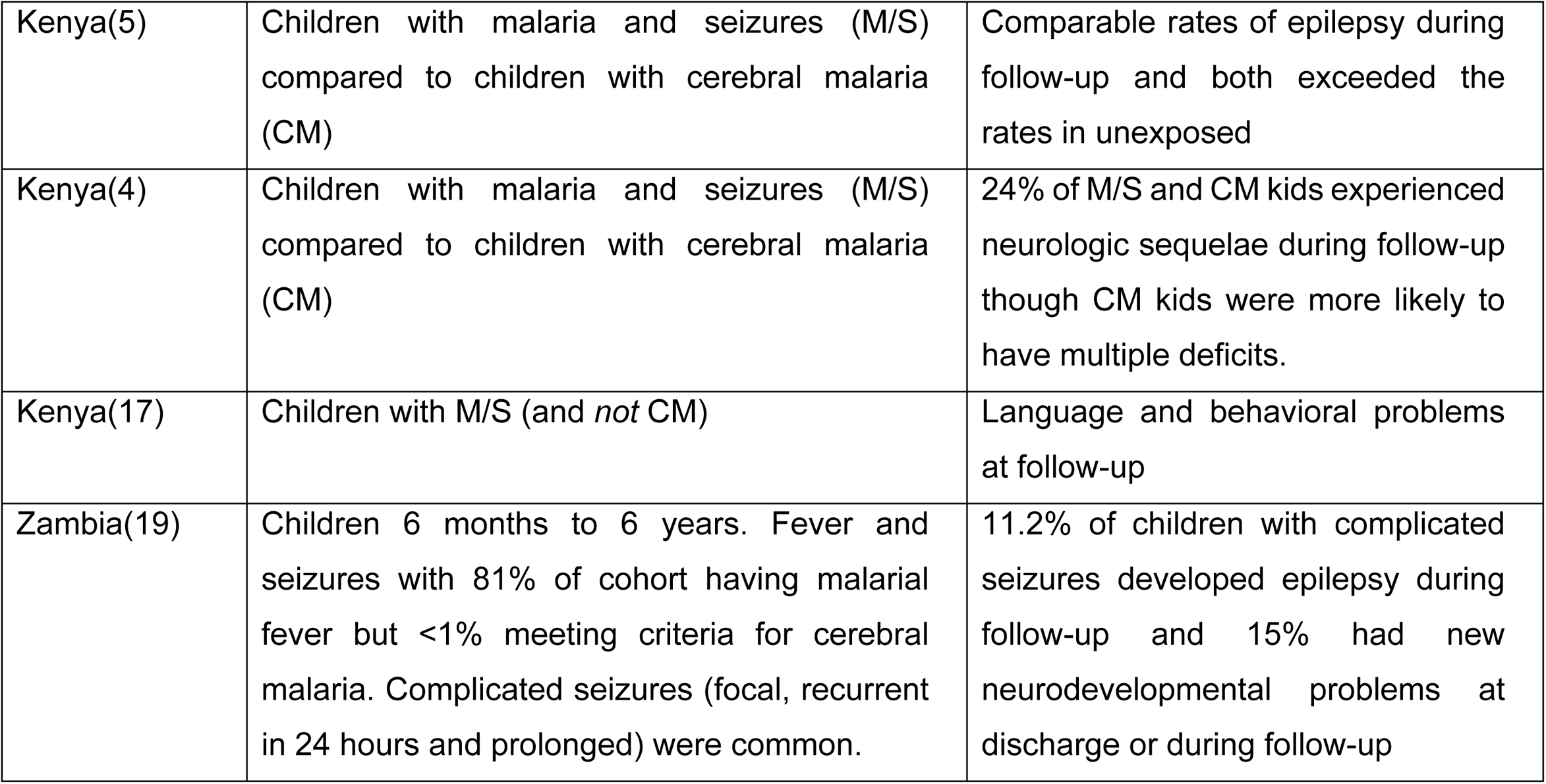
A Review of Neurologic Outcomes after CNS Malaria

###### Fever as a Risk Factor for Malaria-Associated Brain Injury (**Scientific Premise**)

In BMPES, acute symptomatic seizures and a higher maximum temperature (Tmax) during the acute infection were associated with a greater risk of post-malaria neurologic sequelae. Children who developed epilepsy had a mean Tmax of 39.4°C vs. 38.5°C in those who did not develop epilepsy (absolute difference of 0.9°C; p=0.01). In addition, a higher Tmax predicted subsequent behavioral disorders (39.2°C vs. 38.7°C; absolute difference 0.5°C; p=0.04). These data are biologically consistent with the substantial body of evidence that fevers contribute to secondary neurologic injury in the setting of stroke, meningitis, anoxia, hypoxia, and trauma (20–24). In the U.S., induced hypothermia is now standard-of-care for post-anoxic(25) and neonatal hypoxic-ischemic encephalopathy(26–29) and fever-reduction is an important aspect of neurocritical care(30–33). Malaria-induced fevers can be extremely high. Fifteen percent of the BMPES cohort experienced at least one temperature of ≥ 40.0°C (104.0°F). The age-related susceptibility to pediatric malaria substantially overlaps with the age-related phenomena of febrile seizures. Malarial seizures are often complex, multifocal, and prolonged with *status epilepticus* being common (2, 19, 34, 35). See Table 2 for potential mechanisms of fever-induced injury in CNS malaria.

**Table 2:**
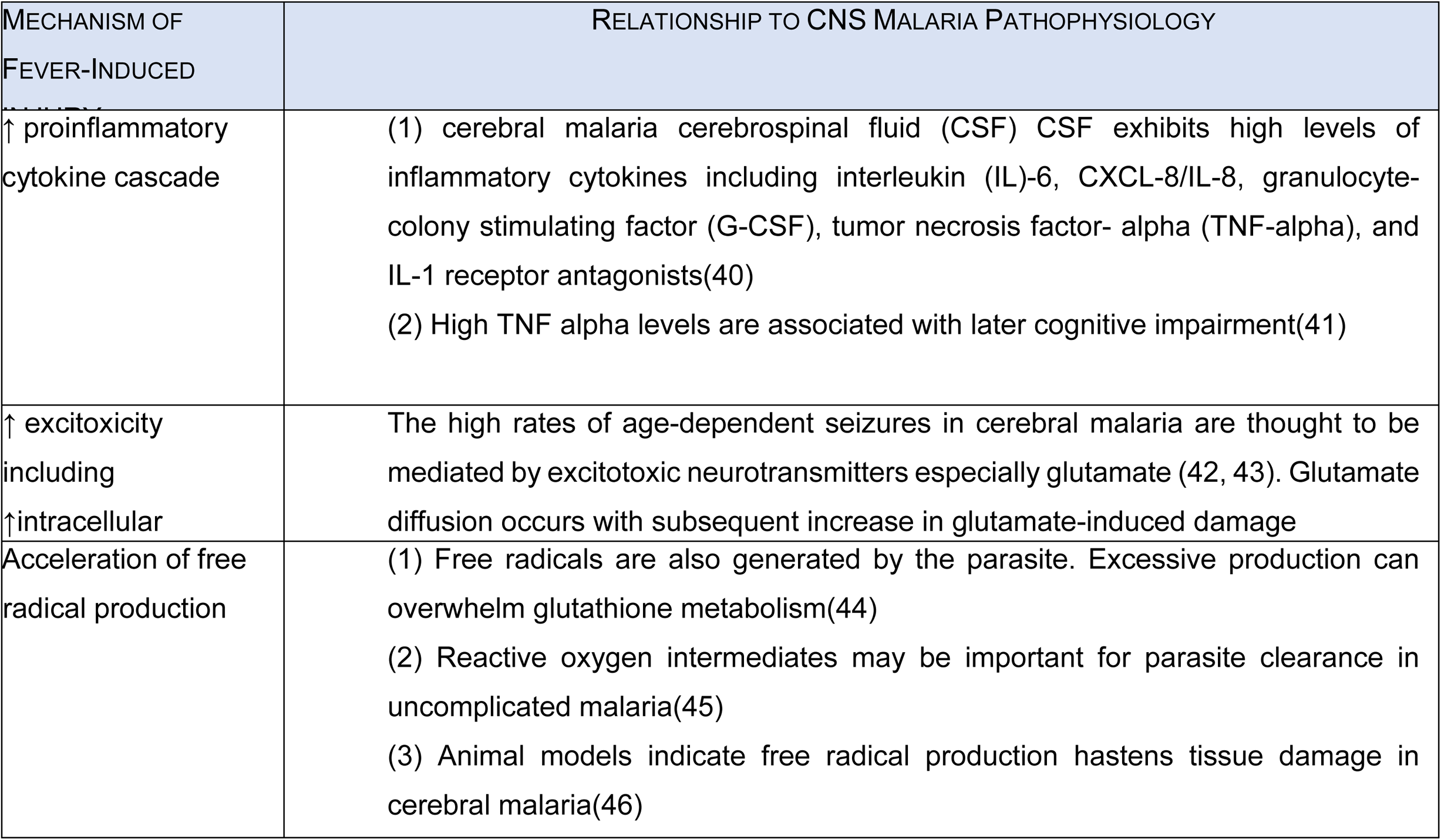
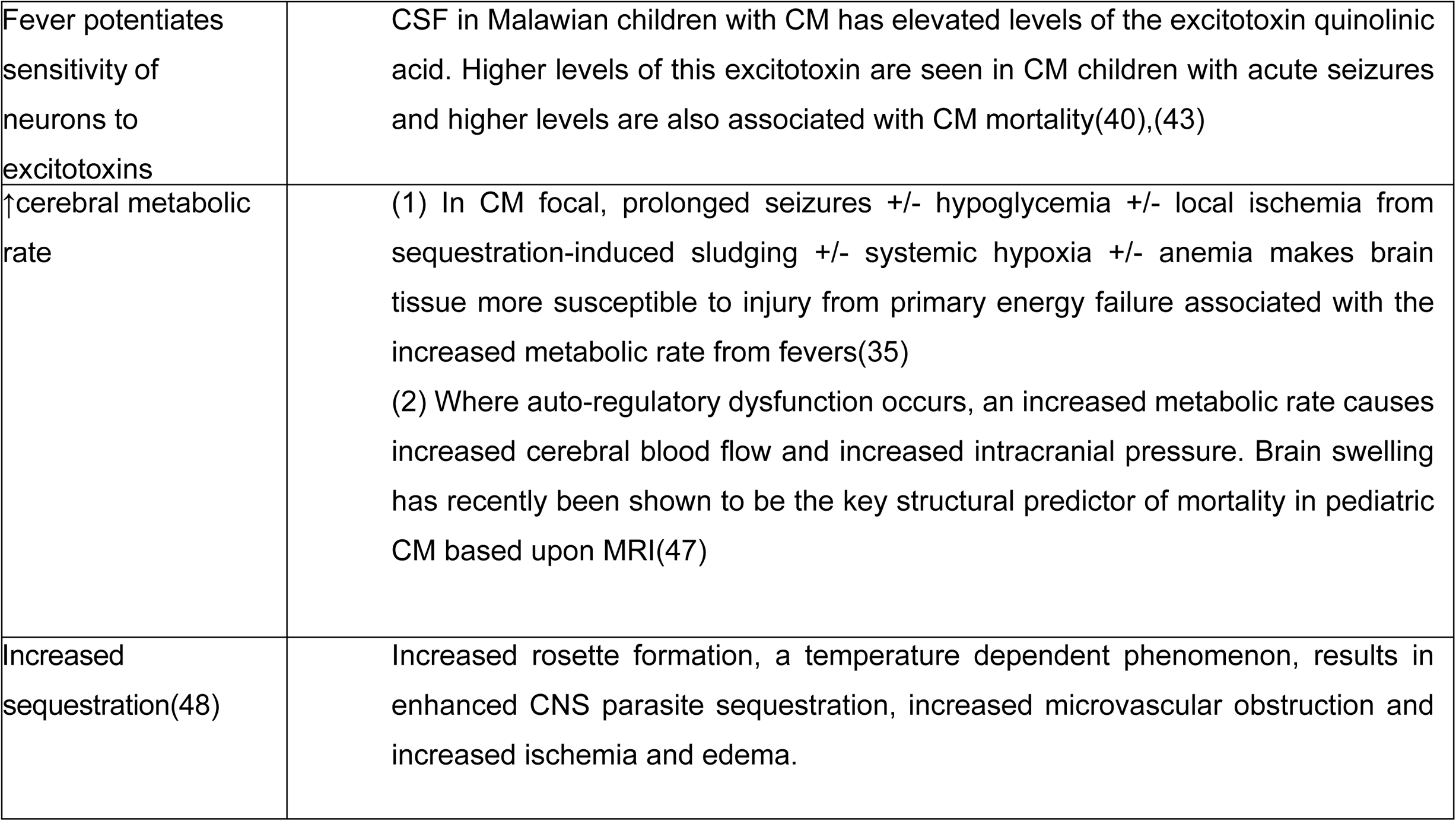
Potential Mechanisms of Fever-Induced Brain Injury in CNS Malaria

###### Standard-of-Care for Malarial Fevers

Despite substantial evidence for the deleterious effects of fever after brain injury, the World Health Organization’s (WHO) Malaria Care Guidelines recommend fever treatment only for rectal T ≥ 38.5°C(36). Since malaria-associated fevers escalate rapidly, treatment based upon this threshold means many children develop significant hyperpyrexia prior to initiating fever-reduction therapy. In the BMPES cohort, among 52 children who were afebrile at the time of admission the mean Tmax during hospitalization was 39.0°C and 13/52 (25%) had a post-admission fever of ≥39.0°C based upon Q2 hourly evaluations. In most malaria care settings, vital signs are only assessed every 6-12 hours, so adherence to WHO Guidelines inevitably result in children with CNS malaria developing and maintaining high fevers for some hours before antipyretics are initiated.

While the neurologist accustomed to aggressive fever management in the neurocritical care unit may find this approach to fever management in pediatric malaria baffling, WHO guidelines and clinical practice patterns are likely predicated upon two factors—first, the well-described dearth of neurologic expertise in malaria-endemic regions(37) and secondly, some evidence suggesting that malarial fevers may be desirable. *In vitro* studies have shown that febrile temperatures inhibit parasite growth with the conclusion being that *“long, high fevers during malaria may be beneficial for parasite clearance*”(38) and a 1997 clinical trial of antipyretics for malarial fevers found that <u>peripheral</u> parasite clearance time was delayed in children who received acetaminophen(39).

###### New Insights into Parasite Behavior and Improved Methods for Measuring Parasite Burden

*In vitro*, uninfected red blood cells (RBCs) stick to those infected with trophozoites and schizonts of *P. falciparum* forming rosettes which are thought to play an important role in sequestration and microvascular obstruction. *S*tudies have shown that rosette formation increases from 37.0 to 40.0°C (48). This temperature-dependent, PfEMP-1 mediated effect doubles RBC cytoadherence and may explain the delayed peripheral parasite clearance time associated with antipyretic use (39). **Essentially, since lowering temperatures may attenuate cytoadherence, peripheral parasite counts may transiently increase as sequestered parasites are liberated into the bloodstream. This may be a desirable effect if by doing so antipyretics improve cerebral blood flow at the capillary-level and reduce sequestration.** Parasitized erythrocytes have also been shown to lose their flexibility at temperatures over 37.0°C especially in the late trophozoite and schizont phase, which likely further exacerbates sequestration (49).

Histidine Rich Protein II (HRP2), a Plasmodium-specific protein in patient plasma, may facilitate quantification of whole-body parasite burden. Recent studies led by Dr. Seydel are supportive of the supposition that sequestered parasites, not those circulating in the peripheral blood, are the primary contributors to severe malaria. They found that mean plasma HRP2 concentrations early in the malaria infection predict progression to CM, even among children receiving treatment(50). Higher plasma levels of HRP2 are also associated with CNS parasite sequestration based upon autopsy findings and bedside retinal assessments with quantitative HRP2 levels of ≥1700ng/ml providing 98% sensitivity and 98% specificity for identifying CNS parasite sequestration (area under the ROC curve 0.98)(51). As such, HRP2 levels provide excellent proxy data for establishing CNS parasite sequestration even in conscious, acutely ill children in whom fundoscopy is problematic.

###### Study Rationale

Studies of other pediatric infections have shown that combination therapy with standard antipyretics can result in reduced fevers, but whether this is true for malaria-associated fevers is unclear. Although antipyretic use with a single agent in one malaria study was associated with prolonged peripheral parasitemia, fever clearance time was not significantly different (32 hours in the antipyretic treated group vs. 43 hours in routine care; p=0.176)(39). Limited fever reduction benefits from single antipyretic agents in malaria have been shown in other studies(52). A randomized controlled trial of acetaminophen (without a loading dose) vs. ibuprofen found an ibuprofen advantage only at 4.5 hours post treatment(53). A controlled trial of acetaminophen vs. placebo for uncomplicated malaria found no differences in mean temperature, but 6/171 children receiving placebo required admission for febrile seizures compared to 2/167 on acetaminophen (54). Antipyretic efficacy has been shown in some studies--acetaminophen is more effective in malaria fever reduction than tepid sponging(55). **No malaria studies to date have evaluated the impact of prophylactic treatment to prevent fevers or interventions using two antipyretics with different mechanisms of action.**

Acetaminophen’s mechanism of action is not fully understood, but it is a potent inhibitor of prostaglandin production in the CNS with minimal anti-inflammatory effects(56). Recent studies suggest that it inhibits the peroxidase functions of COX-1 and COX-2 with selectivity for the inhibition of prostaglandin synthesis (57). In children with non-malarial fevers and a rectal temperature of >39.0°C, 15 mg/kg of acetaminophen resulted in a decrease of 1.07-1.98°C(58), which is greater than the temperature difference associated with better neurological outcomes in the BMPES study. The primary safety concern with acetaminophen use is the hepatotoxicity that can occur when the recommended dosage is exceeded. Ibuprofen is a non-selective COX inhibitor, inhibiting both COX-1 and COX-2. The analgesic, antipyretic, and anti-inflammatory activity of ibuprofen is achieved mainly through inhibition of COX-2, whereas inhibition of COX-1 impacts platelet aggregation and likely mediates gastrointestinal side effects (59). By 1991, some estimates indicated that over 250 million children had received ibuprofen with very few serious adverse events reported (60, 61). The most common ibuprofen side effects include gastritis, impaired platelet function and decreased urinary excretion of sodium. Among non-steroidal anti-inflammatories, ibuprofen is the least likely to cause gastrointestinal bleeding. Studies have shown both acetaminophen and ibuprofen to be safe in children when used individually(62, 63) and in combination(64). Both agents have been shown to be generally safe and effective for fever reduction in children (65–67). Studies in children with non-malarial fevers have found some additional fever-reduction benefit when the two drugs are used together(64). A Malawi-based study of children with uncomplicated malaria comparing acetaminophen and ibuprofen found no difference in fever resolution time and no serious adverse effects from either treatment were identified(68). Acetaminophen and ibuprofen are already approved for fever reduction in the dosage outlined for this planned intervention for use in children aged 2-11 years by the appropriate regulatory agencies (the Pharmacy, Medicines and Poisons Board in Malawi and Zambia Medicines Regulatory Authority) as well as the US Food and Drug Administration.

The rationale for this clinical trial is based upon (1) unmet clinical need in terms of the imperative to decrease the neurologic injury and sequelae that occur in ~250,000 malaria survivors each year, (2) previous epidemiologic data indicating that higher Tmax is associated with neurologic sequelae in CNS malaria, (3) the very plausible biological mechanisms of malarial fever-related secondary brain injury detailed in Table 2, (4) *in vivo* and *in vitro* data on temperature dependent parasite behavior and whole body parasite burden suggesting that findings from previous research linking fever reduction to slowed parasite clearance may be erroneous.

###### Capacity to Conduct Clinical Trials and Capture Delayed Outcomes

Our prior outcome studies of severe malaria (2, 19) has provided us with the experience of (successfully) negotiating the regulatory requirements and compliance challenges of conducting an NIH-funded clinical trial in the African setting has provided us with the experience of (successfully) negotiating the regulatory requirements and compliance challenges of conducting an NIH-funded clinical trial in the African setting.

###### Further Insights into the Role Fever Plays in the Pathophysiology of Neurologic Injury in CNS Malaria

Is fever independently associated with adverse neurologic outcomes? Children with CNS malaria are of an age to be susceptible to febrile seizures and acute malarial seizures are a well-established risk factor for neurologic sequelae in malaria survivors. To evaluate whether temperature independently affects neurologic outcomes in CNS malaria, we used the BMPES data to examine the relationship between Tmax and adverse neurologic outcomes in a regression model that adjusted for the presence/absence of seizures during the acute admission. **After controlling for acute seizures, a higher Tmax remained associated with adverse neurologic sequelae during follow-up (p=0.029)**.

does the relationship between fever and adverse neurologic outcome hold true for children with cns malaria more broadly defined? The BMPES study, which identified a higher Tmax as being a risk factor for neurologic sequelae in CNS malaria, only included children with retinopathy positive CM(2). Although children with the more broadly defined CNS malaria experience similar rates of neurologic injury evident after recovery (detailed in Table 1 above), little is known or published about the impact of fever on outcomes in this broader population. To examine the effects of fever on a broader population of children with malaria, we compared Tmax in retinopathy negative children with and without neurologic sequelae who were enrolled during the BMPES study. The clinical characteristics(69), neurologic outcomes(70) and MRI findings(71) of these retinopathy negative children have been previously described. Among 35 retinopathy negative children, the mean Tmax in children with neurologic sequelae was 39.2°C vs. 38.7°C in those without sequelae—a finding very consistent with the difference in mean Tmax found in the retinopathy positive BMPES cohort suggesting that fever likely plays a role in neurologic injury across the broader spectrum of CNS malaria.

what is the pathophysiologic basis for fever-mediated secondary brain injury? HOW CAN WE BEST CAPTURE THIS EXPOSURE? Based upon numerous epidemiological studies (20–24) (25–29)(30–33)(30–33)(72) data from malaria experiments conducted by White et al. led them to conclude that keeping the temperature at 37.0°C will delay cytoadherence by 3 hours and allow up to 25% of the ring parasites to be removed before sequestration (48). Fortunately, the 0.5°C difference in mean Tmax in children with vs. without sequelae in the BMPES study does offer some insights into what constitutes a clinically meaningful difference in Tmax. To further explore whether Tmax is the most sensitive measure for predicting fever-associated neurologic sequelae from malaria, we returned to the BMPES study data and examined the association between adverse neurologic outcome and area under the fever-time curve based upon Q2 hourly assessments using a fever threshold of 37.0°C and a threshold of 37.5°C. Unlike Tmax, these alternate measures of temperature exposure were not associated with adverse neurologic outcomes, so our primary outcome remains Tmax. Unlike Tmax, these alternate measures of temperature exposure were not associated with adverse neurologic outcomes, so our primary outcome remains Tmax.

#### HRP2 Studies

**Figure 1:**
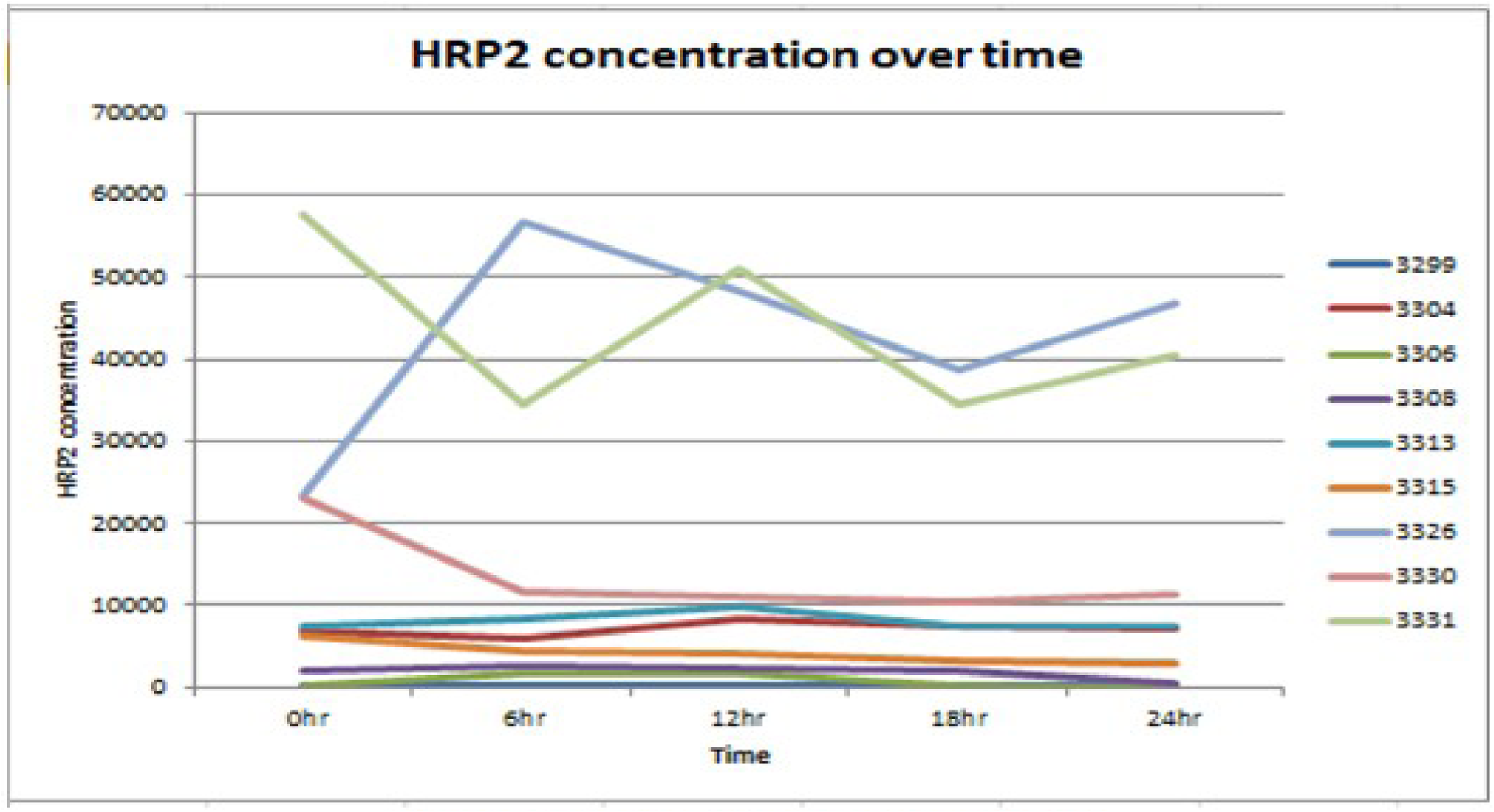
HRP2 Pharmacokinetics (n=9)

Our studies of HRP2 levels in Malawian children have supported the importance of quantifying the parasite burden through means other than a peripheral blood smear. HRP2 levels in the periphery are affected by multiple variables, including level of sequestration and rate of parasite clearance post-treatment. In a preliminary experiment we measured Q6 hour HRP2 levels post-treatment and found that marked differences occur despite identical treatment. We therefore will measure HRP2 levels every 6 hours in both arms of the trial with the hypothesis that in the aggressive antipyretic arm sequestration will be reversed and thus area under the concentration-time curve of HRP2 levels will decrease

## Objectives

### Hypothesis

*We hypothesize* that children who receive aggressive antipyretic therapy will have:

A lower mean Tmax using continuous temperature monitoring

(ii) Decreased acute symptomatic seizures during the 72-hour treatment period
(iii) A lower incidence of long-term neurologic sequelae. This will be assessed in the more definitive study to follow if we meet our “go” criterion—meaning we reject the null hypothesis of no treatment effect with findings of a lower mean Tmax in the aggressive antipyretic treatment group compared to usual care.

### Primary objectives

To conduct a randomized, double-blind, placebo-controlled trial of aggressive antipyretic therapy to reduce Tmax in pediatric CNS malaria and evaluate the “proof-of-concept” for potential neuroprotective efficacy. Children admitted with CNS malaria, *defined as malaria with impaired consciousness and/or seizures*, will be randomized to usual care (15 mg/kg acetaminophen Q6 hours as needed for ≥ 38.5°C) vs. aggressive antipyretic therapy with prophylactic acetaminophen (30 mg/kg load followed by 15 mg/kg Q6 hours) PLUS ibuprofen (10 mg/kg Q6 hours) from time 0 to hour 72, the timeframe in which most children with malaria will defervesce

### Secondary objectives

a. To evaluate the safety of aggressive antipyretic therapy. After enrollment, serial assessments to identify incident hypoglycemia, bleeding/clotting problems, renal insufficiency, and/or hepatic dysfunction will be undertaken. Local Study Monitors will review adverse events for individual participants in real time. Interim analyses will be conducted annually with explicit stopping rules based upon mortality.
b. To assess the effect of aggressive antipyretic therapy on parasite sequestration and parasite clearance, we will evaluate host parasite burden through Q6 hourly quantitative parasite counts as well as HRP2 quantitation. *We hypothesize* that fever treatment will reduce sequestration resulting in lower HRP2 levels based upon area under the curve during the 72-hour treatment period and that an associated transient paradoxical increase in peripheral parasitemia with a higher proportion of older parasites will be seen as centrally sequestered parasitized red blood cells are released.

## Trial design

The trial will be randomized, double-blind, placebo-controlled trial of aggressive antipyretic therapy with the primary outcome being the maximum temperature occurring in the 72 hours after presentation with acute malaria. Children with malaria and neurologic symptomatology (either cerebral malaria or CNS malaria) will be randomized to receive aggressive antipyretic therapy with scheduled acetaminophen starting with a 30mg/kg load following by 15mg/kg Q6 hourly PLUS ibuprofen 15mg/kg Q6 hourly vs. usual care. Usual care is 15mg/kg of acetaminophen Q6 hourly as needed for a temperature of ≥38.5°C. A total of 284 children (142 in each study arm) will be enrolled.

## Methods: Participants, interventions and outcomes

### Study setting

The study will be carried out in three settings: the Pediatric Research Ward (PRW) at Queen Elizabeth Central Hospital in Blantyre, Malawi; the Pediatric Intensive Care Ward (PICU) at University Teaching Hospital in Lusaka, Zambia; the High Acuity Section within the Pediatric Ward at Chipata Central Hospital, Chipata, Zambia.

### Eligibility criteria

Parents or guardians must provide written informed consent in their local language prior to any procedures or treatments taking place.

#### Inclusion Criteria

- Age 2-11 years (24 to 132 months)
- Evidence of *P. falciparum* malaria infection by peripheral blood smear or rapid diagnostic test
- CNS symptoms associated with malaria. CEREBRAL MALARIA: Impaired consciousness with a Blantyre Coma Score (BCS)(73) ≤2 in children under 5 years or a Glasgow Coma score (GCS) ≤10 in children ≥5 years **OR** CNS MALARIA: Complicated seizure(s), meaning prolonged (>15 minutes), focal or multiple; or impaired consciousness or other evidence of impaired consciousness (confusion, delirium) without frank coma (BCS>2, GCS =11-14)

#### Exclusion Criteria

- Circulatory failure (cold extremities, capillary refill > 3 seconds, sunken eyes, ↓ skin turgor)
- Vomiting in the past 2 hours
- Serum Cr > 1.2 mg/dL
- A history of liver disease
- Jaundice or a total bilirubin of >3.0mg/dL
- A history of gastric ulcers or gastrointestinal bleeding
- A history of thrombocytopenia or other primary hematologic disorder
- Petechiae or other clinical indications of bleeding abnormalities
- A known allergy to ibuprofen, acetaminophen, aspirin or any non-steroidal medication
- Any contraindication for nasogastric tube (NGT) placement and/or delivery of enteral medications.

### Who will take informed consent?

Staff who consent study subjects are local healthcare workers fluent in the local language.

## Interventions

### a. Study Intervention

i. Intervention arm-scheduled dosing every 6 hours with acetaminophen (30mg/kg p.o. load then 15mg/kg Q6 hours) and ibuprofen (10mg/kg Q6 hourly), starting at enrollment (hour 00) and continuing through hour 72 including a final dose at hour 72.
ii. Usual care arm-acetaminophen 15mg/kg Q6 hourly as needed for temperature ≥ 38.5°C based upon axillary temperatures taken in the course of routine clinical care (starting at enrollment (hour 00) and continuing through hour 72 including a final dose at hour 72)
iii. Children who have received acetaminophen in the 24 hours prior to presentation, based upon the available medical record or report from the parent or guardian, will not be given a loading dose of acetaminophen. Instead, they will be commenced on the maintenance dose of 15mg/kg Q6 hourly.

### b. Criteria for discontinuing or modifying allocated interventions

Criteria for discontinuing or altering interventions are listed in Table 3 below.

**Table 3.**
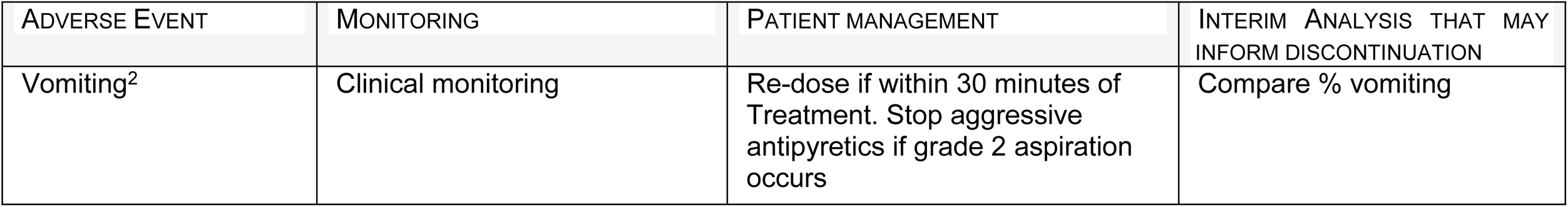
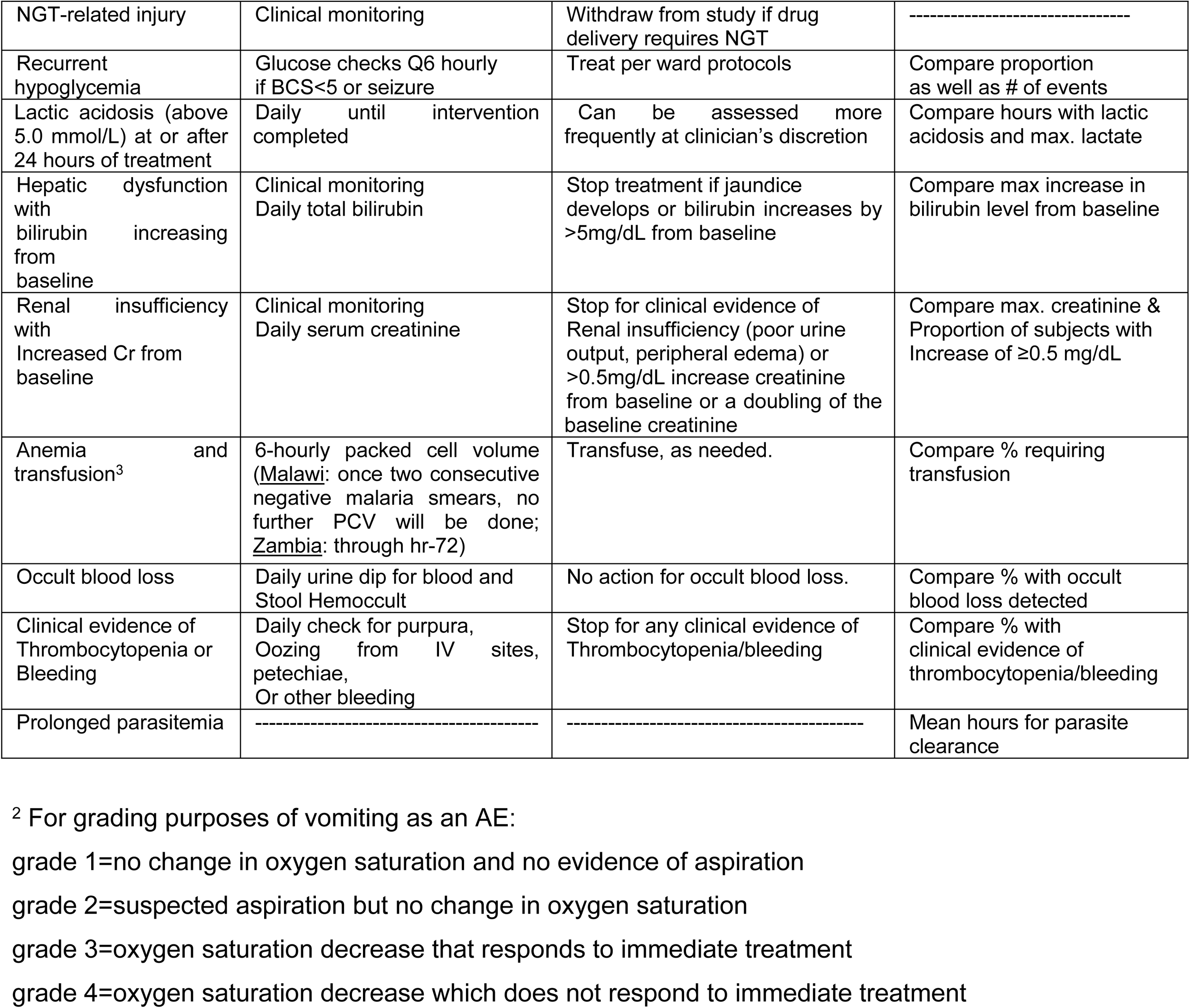
Plan for Monitoring and Discontinuation

Children will be withdrawn from the study if requested by a parent or clinician caring for the patient or recommended by the Local Study Monitor (LSM) or DSMB

### c. Strategies to improve adherence to interventions

All doses will be directly administered by a study trained nurse. The administering ward nurse will directly observe the oral intake of the medication and document oral adherence. If vomiting occurs with 30 minutes of oral administration, a repeat dose will be given, and documentation of the event made.

### d. Relevant concomitant care permitted or prohibited during the trial

#### Concomitant Treatment and Investigations

All treatments routinely used in the acute care of children with severe malaria will be provided (75). These include antimalarials, anticonvulsants, and blood transfusions. The duration and characteristics of all clinical seizures are documented in a bedside “fitting chart”. Due to the significant risk of subclinical seizures in comatose children with malaria (2), a daily, routine, 30-minute EEG will be completed in any child with a BCS≤4 or GCS≤10. When necessary, nasogastric tube (NGT) placement will be undertaken by experienced ward nurses. The NGT will be removed and oral administration resumed as soon as the child is able to swallow.

### Provisions for post-trial care

If clinically indicated, follow-up evaluations and care will be provided by the admitting service. Patients that are enrolled into the study are covered by indemnity for negligent harm through an indemnity arrangement arranged at each study site. Insurance to cover for non-negligent harm associated with the protocol will be procured prior to enrollment. This will include cover for additional health care, compensation or damages whether awarded voluntarily by the Sponsor, or by claims pursued through the courts.

### Outcomes

#### Primary outcome measures

Maximum temperature (T_MAX_). T_MAX_ will be defined as the highest temperature during the study duration (72 hours) in degrees Celsius recorded by a continuous temperature monitor. Regardless of treatment allocation, temperature will be continuously assessed for 72 hours using a non-invasive, accurate temperature monitoring system that continuously measures patient temperature using an external temperature sensor placed in the axilla. The continuous temperature display will be concealed from nursing staff responsible for measuring temperatures for clinical care and delivering rescue fever treatment based upon the protocol. Temperature data will be captured locally on a device and transmitted via email to a dedicated study account.

The continuous monitoring device is not MR compatible. If Tmax is a clinical temperature obtained when continuous monitoring data was not available, the clinical Tmax will be used as the primary outcome.

#### Secondary outcome measures

a. Seizures will be assessed for occurrence and severity, from t=0 to t=72 hours based upon “Fitting Charts” and a daily routine EEG in comatose (BCS≤4 and GCS≤10) children. The daily EEG will be reviewed by the neurologist covering the PRW or PICU Neurodiagnostic Service. **Seizure outcome has three categories—no seizures, a single brief (<15 minute) seizure, and multiple or prolonged (>15 minutes) seizures.** These seizure categories are clinically relevant and correspond to risk factors for subsequent neurologic sequelae in our previous Zambia-based prospective cohort study of fevers and seizures
b. HRP2 (i.e., total body parasite burden) is the quantitative level reported as ng/ml in plasma. Samples for HRP2 determination will be obtained Q6 hourly via finger prick. Levels will be determined by ELISA and will not be available in real time.
c. QUANTITATIVE PARASITE COUNT (qPC, peripheral parasite burden) is the number of parasites per microliter of blood based upon standard laboratory measures and obtained Q6 hourly via finger prick. Samples will cease to be collected when no parasites are seen in two subsequent, consecutive samples at the Malawi site; in Zambia, malaria smears will be obtained Q6 hourly through each participant’s 72 hours of enrollment but then read at a later time by a Malawian microscopist. For enrollment and clinical care considerations at the Zambia site, malaria rapid diagnostic tests (RDTs) will also be performed to guide anti-malarial treatment decision-making. Values will be obtained by counting the proportion of 500 RBCs that are parasitized under high-power microscopic evaluation of a thin blood smear. The PCV will also be obtained at each time point and will be used to estimate RBC count using the formula PCV/8 = RBC X 10^6 per microliter of blood. Estimated RBC count will be multiplied by the percent RBCs parasitized to arrive at the number of parasites per microliter of blood
d. hemozoin will be assessed as the percentage of parasites with hemozoin deposition based upon 100 parasites viewed in a high-powered field at each qPC
e. exposure as measures by the area under the curve (AUC) for 72 hours for fever ≥38.5°C

#### Additional demographic and clinical data to be collected

- Gender
- Age
- Seizure history
- Admission temperature
- Difficult birth history
- Developmental status as assessed by Ten Question screen
- Family history of seizures
- Retinopathy status in comatose subjects.

### Participant timeline

**Table 4:**
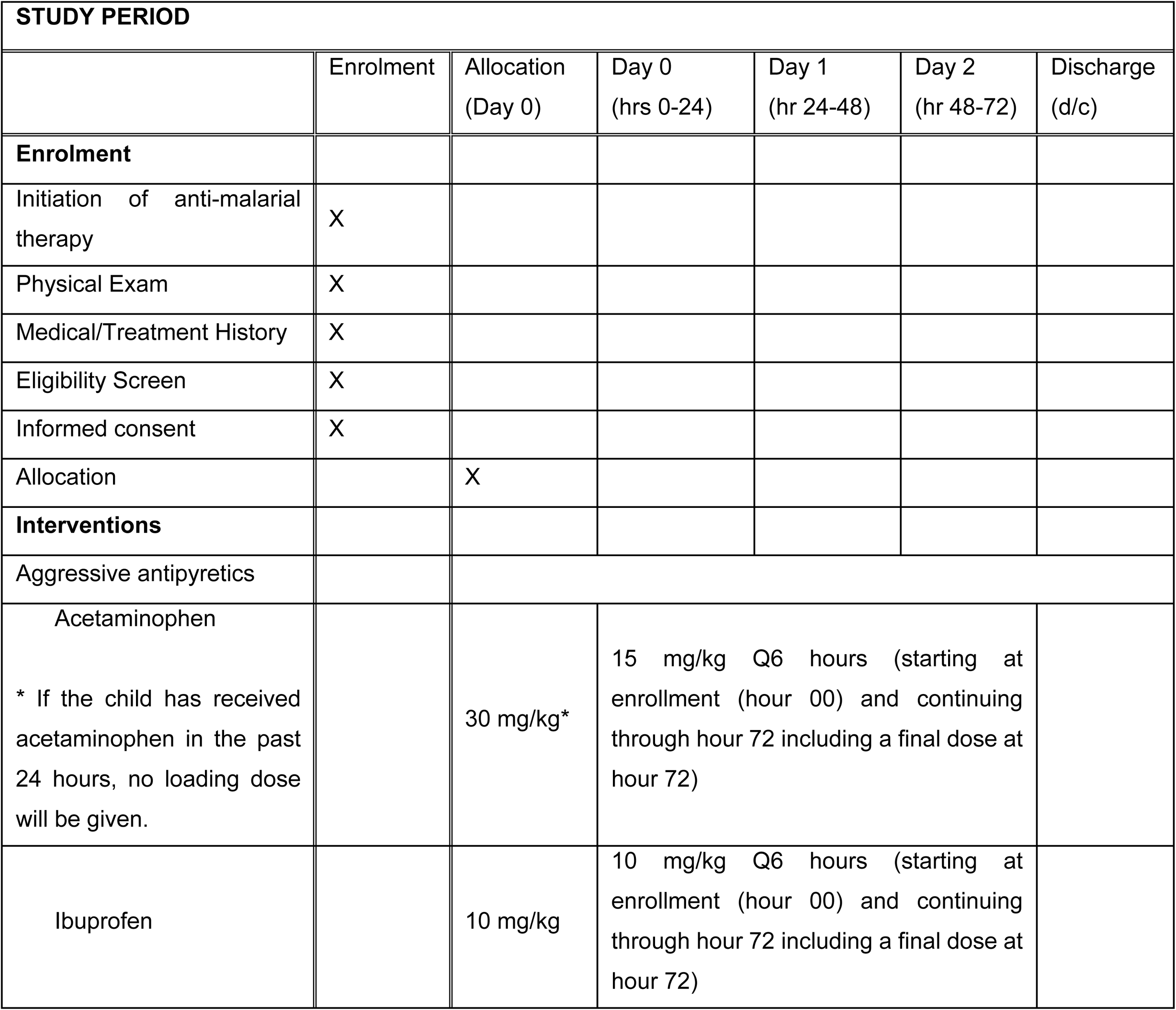
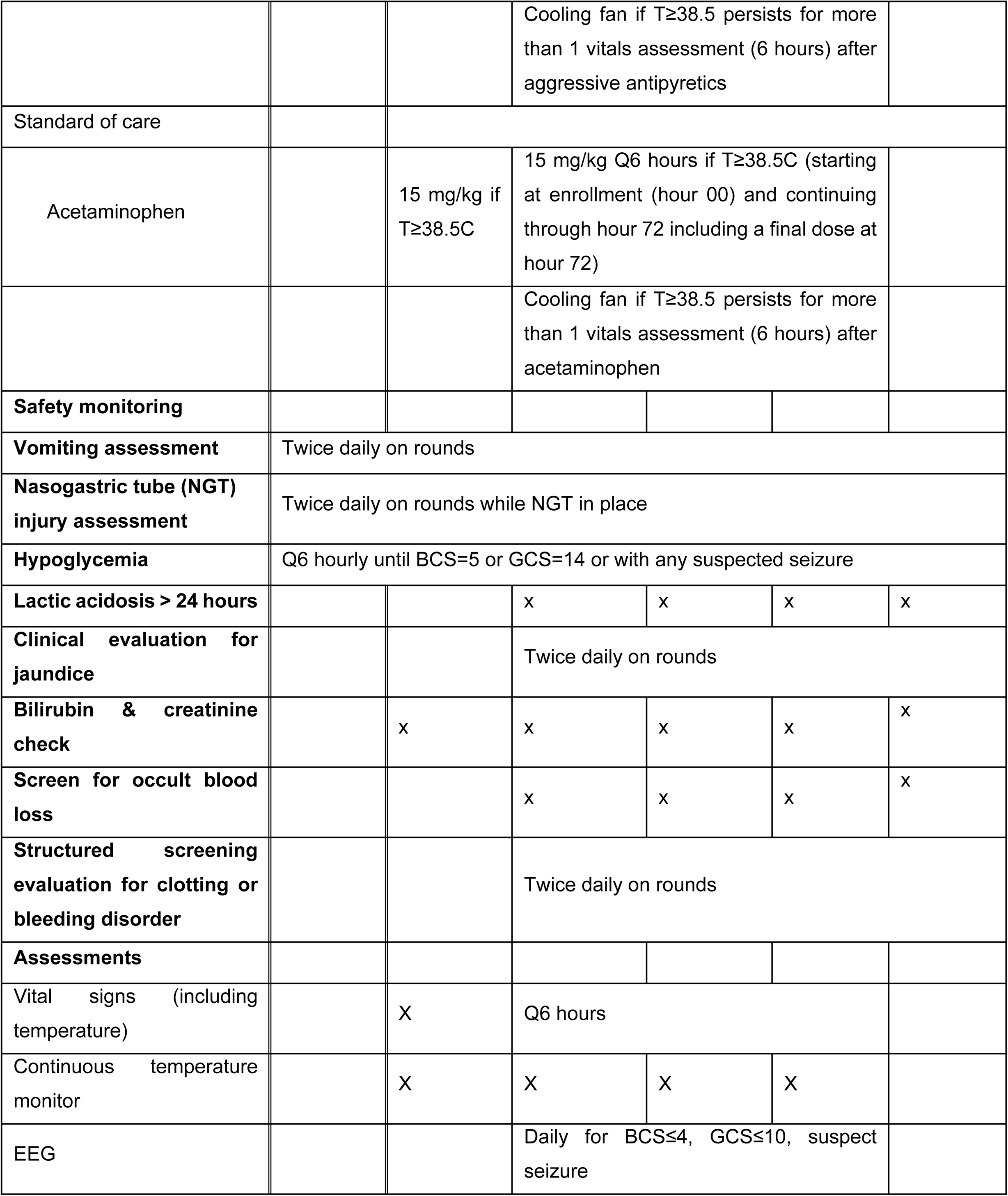
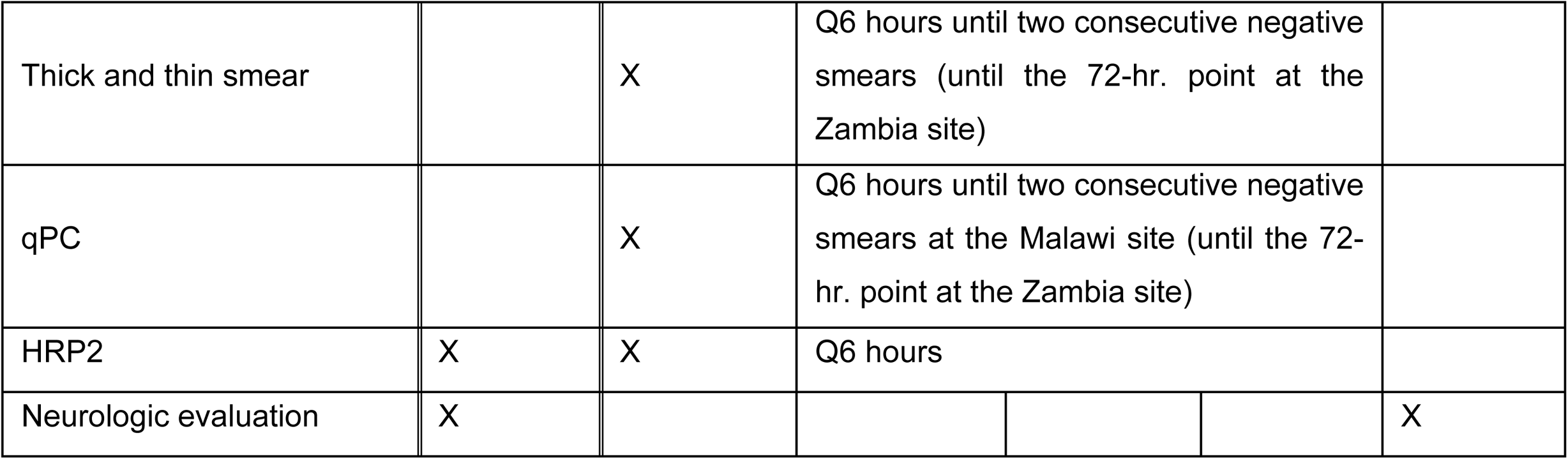
Participant Timeline

### Sample size

The primary analysis in this study will involve a comparison of the adjusted mean Tmax between the treatment groups. In the BMPES study, the difference in mean Tmax for children with vs. without behavioral disorders was 0.5°C, and it was 0.9°C between those with and without epilepsy (2). A receiver operating characteristic curve analysis suggests that a good balance between sensitivity and specificity for Tmax in predicting epilepsy is achieved for a Tmax of 38.9°C and the mean Tmax for children who experienced epilepsy was 39.4°C suggesting that a reduction in temperature of 0.5°C is a clinically meaningful reduction. CM at the proposed study sites has a mortality rate of ~15% and most children who die will do so within 12-24 hours after admission and are unlikely to benefit from fever reduction measures. The effect of mortality on the sample size is manifested through attenuation of the group difference in mean Tmax of 0.5 °C. Under the assumption that mortality is comparable in the two groups and that there is no treatment effect in those who die, the attenuated treatment effect will be (0.5 × 0.85) + (0.0 × 0.15) = 0.425 °C. Assuming a standard deviation of the maximum temperature of 1.1 °C as was present in the BMPES study population, **a sample size of 142 children per group will provide 90% power to detect a group difference in mean Tmax of 0.425°C, using a two-sample t-test and a 5% significance level (two-tailed).** The 0.5°C threshold (before attenuation) is based on our observational data that adverse neurologic outcomes in cerebral malaria occur at mean Tmax differences of 0.5°C (2).

For the outcome of acute symptomatic seizures, data collected during past malaria studies (2, 19) suggest that the incidence of seizures after admission will be 40% in children with CM and 24% in CNS malaria. Assuming an overall seizure incidence after enrollment of 32% in the usual care group (68% with no seizures, 25% with a single brief seizure, and 7% with multiple or prolonged seizures), a sample size of 142 children per group will provide 80% power to detect a shift in the aggressive antipyretic treatment group to 82% with no seizures, 15% with a single brief seizure, and 5% with multiple or prolonged seizures, representing a common log-odds ratio of 0.7656. The method due to Whitehead(76) was used for this calculation.

Since we are preferentially recruiting to ensure that approximately 50% of children enrolled have CM and past experience indicates that 70% of CM admissions during the rainy season (i.e., malaria season) are retinopathy positive, we anticipate that there will be 100 children included in the secondary analysis of Tmax differences between treatment groups among children with CM and CNS parasite sequestration compared to all others. Assuming a standard deviation of Tmax of 1.1 °C(2), **a sample size of 50 children per group will provide 80% power to detect a group difference in mean Tmax of 0.62°C (after attenuation due to mortality), using a two-sample t-test and a 5% significance level (two-tailed).** Sample size considerations cannot be provided for Hypothesis 2 due to the absence of preliminary data on HRP2 levels, including the absence of any reasonable data on standard deviation for HRP2 levels or the percentage of mature parasites.

### Recruitment

#### Blantyre (Malawi)

Recruitment will be directly from the Accident and Emergency Department where study nurses knowledgeable of the inclusion criteria will be based. Patients will be directly transferred to the Pediatric Research Ward (PRW). The PRW serves as the de facto intensive care unit for the hospital, hence many of the eligible patients will be transferred to the PRW independent of this trial

#### Lusaka (Zambia)

Recruitment will be directly from the Emergency Department where a study nurse knowledgeable of the inclusion criteria will be based. Patients will be transferred to the Pediatric Intensive Care Unit (PICU) or the Admission Ward beds adjacent to the PICU depending upon medical need.

#### Chipata (Malawi)

Recruitment will be directly from the Emergency Department where study staff knowledgeable of the inclusion criteria will be based. Patients will be transferred to the High Acuity section within the Pediatric Ward.

## Assignment of interventions: allocation

### Sequence generation

The randomization process is designed to be blinded to data collectors. The randomization plan will be developed by the unmasked programmer at UR Biostatistics. It will be generated using a SAS program in accordance with Biostatistics standard operating procedures. Subjects will be randomly assigned with equal allocation to the two treatment groups. The randomization will be stratified by country and will include blocking to ensure approximately equal allocation of the treatments within each stratum after a specified number of subjects have been enrolled in that stratum. The randomized allocation will be provided to the compounding and study site pharmacists preparing the intervention and placebos.

### Concealment mechanism

Patients will be assigned to treatment groups using blocked randomization with randomly selected block sizes. Randomization will delineate treatment allocation to sequential, previously delineated study IDs for each site. Details of random allocation will be provided to the local study pharmacist for preparation of treatment packages by the US-based compounding pharmacist as received from the unmasked UR Biostatistics programmer. In addition, sealed opaque envelopes with treatment allocation by study ID will be provided to the study sites in case there is the need to break the code for an individual patient. At the completion of the study, all envelopes will be returned to UR Biostatistics.

### Implementation

The unmasked biostatistician from the University of Rochester will generate the allocation sequence. Research staff who are also healthcare providers at each facility will enrol participants who will receive sequentially numbered study drug treatment packets prepared by the study pharmacists based upon allocation instructions from the unmasked biostatistician.

## Assignment of interventions: Blinding

### Who will be blinded

Allocation will be concealed from all staff. Continuous temperature readings will be captured electronically but the display will be concealed from nursing staff who provide treatment for interim fevers based upon treatment protocol and temperature measured clinically per usual care on the ward. The EEG interpreter will be blinded to allocation and temperature.

### Procedure for unblinding if needed

Sealed envelopes indicating allocation will be provided at the study sites if unblinding is needed for patient care.

## Data collection and management

### Collection of data

a. Temperature and Tmax

i. Temperature: Temperature for clinical care purposes will be acquired from the vital sign assessments performed by the study nurses.
ii. Tmax for primary study outcome will be acquired from the continuous temperature monitoring device. This data will not be visible to or made available to study nurses providing clinical care
b. Fever Exposure. Fever exposure is the area under the curve (AUC) for 72 hours for fever ≥ 38.5°C and this will be uploaded into the REDCap database from the CRF. The raw data captured every 2 minutes will also be captured in an Excel spreadsheet to then be emailed to the biostatistician separately.
c. Seizure activity. Seizure activity will be recorded by the study clinicians on a fitting chart attached to each subject file. This will capture the time, nature, and treatment of each episode of seizure activity. Subjects will also undergo a routine 30-minute 21-lead EEG every day while BCS≤ 4 or GCS≤10. Seizure activity on EEG adhere to International League against Epilepsy (ILAE) guidelines for electrographic seizure
d. qPC. qPC will be determined by standard protocol as delineated in the SOP. Briefly the number of parasites in 500 RBCs will be determined by microscopy. This will be translated to parasites per microliter using the PCV divided by eight as an estimate of RBC X 10^6 per microliter (ul).
e. HRP2. HRP2 concentration per ul will be determined by previously published methods (51). Briefly, plasma will be obtained from the microhematocrit tubes used for PCV determination. After determination of PCV (if clinically needed), tubes will be scored at the RBC/plasma interface with a triangular file and broken. A narrow pipette tip will be inserted in the plasma containing portion and the plasma removed. Plasma will be transferred to a bar code-labeled microcentrifuge tube and stored at −20°C until analysis. At the time of analysis, plasma diluted at a ratio of 1:500 in phosphate buffered saline (PBS) as well as a titration of a stock of recombinant HRP2, will be plated onto a plate precoated with anti-HRP2 antibody (Cellabs, Brookvale, Australia). The manufacturer’s protocol will then be followed, except for the modification of all incubations being carried out at 37°C in a humidified chamber. In brief, a 1-hour sample incubation step will be followed with extensive washing with PBS/0.1% Tween. One hundred microliters of the second conjugated antibody will be plated and allowed to incubate for 1 hour. The conjugate will subsequently be washed off and 100 μL of substrate added for 15 minutes, during which color change will be observed. This reaction will be stopped with 1N HCl and the plate analyzed at optical density (OD) 450. A standard curve will be generated from the recombinant protein and the readings from the diluted unknown samples will be compared to this curve to calculate HRP2 levels. If the initial dilution of the sample results in an OD reading that does not lie within the linear range of the recombinant protein, the sample will be rediluted at either 1:1000 or 1:100 to result in OD values within the dynamic range of the ELISA. Samples that remained off scale low, despite redilution, will be assigned a value equivalent to the lowest possible detection value of the ELISA.
f. EEG. EEGs will be obtained on the day of admission and daily thereafter until recovery (i.e., if BCS≤4, GCS≤10). EEGs will be obtained using a modified 10-20 system, and meet the American Electroencephalography Society’s guidelines for EEG (77). For clinical purposes EEGs will be systematically reviewed by the neurophysiologist providing clinical support for the respective site the day the EEG is obtained.
g. Data collection Process Data will be collected on paper-based forms included in the subjects’ clinical files during the admission, and study-specific variables will be captured in these forms by study personnel. A paper-based data abstraction to a case-report form will then be completed by local study personnel before double data entry into REDCap.

### Retention of subjects

The study period is limited to 72 hours and >90% of children admitted with CNS malaria are inpatient for 72 hours or longer. To encourage retention among those ready for discharge before the study completion, transportation home and a meal allowance will be provided for their parent(s). The informed consent process will also be very explicit and clear about the need for at least a 72-hour admission and children with parents unwilling to commit to this will not be enrolled. If clinical or safety concerns are present at discharge, a 30-day follow-up will be scheduled as per clinical care protocol for both study sites.

### Data management

The REDCap system is a secure, web-based application that is flexible enough to be used for a variety of types of research. It provides an intuitive interface for users to enter data and real-time validation rules (with automated data type and range checks) at the time of data entry. REDCap offers easy data manipulation with audit trails and functionality for reporting, monitoring and querying patient records, as well as an automated export mechanism to common statistical packages (SPSS, SAS, Stata, R/S-Plus). The Double Data entry feature allows a project administrator to designate two users per study to perform double data entry. Other users are then able to compare the data entry results, using the data comparison tool. This requires that the Double Data Entry feature be activated for the study.

Through the University of Rochester Medical Center’s Center for Health and Technology (CHeT), the Clinical Translational Science Institute (CTSI) Informatics Core will serve as a central facilitator for data processing and management. REDCap data collection projects rely on a thorough, study-specific data dictionary defined in an iterative self-documenting process by all members of the research team, with planning assistance from the CTSI Informatics Core. CHeT’s iterative development and testing processes results in a well-planned data collection strategy for individual studies. REDCap servers are housed in a local data center at the University of Rochester and all web-based information transmission is encrypted. REDCap was developed in a manner consistent with HIPAA security requirements and is recommended to University of Rochester researchers by the URMC Research Privacy Officer and Office for Human Subject Protection. Access to data in the REDCap application and database will be controlled by using user definitions and permissions. The PI will have access to all study data and will direct access for all users. Audit logs are created automatically within the REDCap application to capture the complete user history of database activity.

Data entry for the subject is expected to be complete within one week of randomization. Most data entry errors will be resolved by the Double Data Entry process operating on local machines at each site. Therefore, Data Management will be primarily responsible for ensuring CRFs are entered in a timely manner and are complete. Reports of delayed data entry and missing CRF elements will be generated weekly for the CHeT project manager to review with sites.

### Confidentiality

All laboratory specimens, evaluation forms, reports, EEGs, and other records that leave the site will be identified only by the study ID number to maintain subject confidentiality. All records will be kept in locked study-dedicated offices. All computer entry and networking programs will be done using SIDs only. Clinical information will not be released without written permission of the subject, except as necessary for monitoring by the involved institutions ethics review boards (RSRB, COMREC, and UNZA BREC), the Malawi Pharmacy Medicines and Poisons Board, the Zambia Medicines Regulatory Authority, the NINDS, the OHRP, the sponsor, or the sponsor’s designee.

## Statistical methods

The analyses presented below will be performed according to the intention-to-treat principle and will include all available data from all randomized subjects. The assumptions underlying all statistical models will be thoroughly checked using graphical and inferential methods and remedial measures will be taken (e.g., transformations, consideration of alternative models) if these assumptions are found to be seriously violated. A significance level of 5% will be used for hypothesis testing.

Analytic Plan for Hypothesis 1(i):

*Children who receive aggressive antipyretic therapy will have lower mean Tmax during the 72-hour treatment period based upon continual temperature assessments*.

The analysis of the Tmax will involve fitting an analysis of covariance model with treatment group as the factor of interest, site and disease severity (CM vs CNS malaria) as stratification factors, and admission temperature as a covariate. The estimated treatment effect (difference in adjusted group means between the aggressive antipyretic therapy and usual care groups) and its associated 95% confidence interval, as well as a t-test for significance of the treatment effect, will be derived from this model.

Analytic Plan for Hypothesis 1(ii):

*Children who receive aggressive antipyretic therapy will be less affected by acute symptomatic seizures during the malaria infection based upon clinical assessments and daily routine EEGs for those with BCS≤4 and GCS≤10*

To compare seizure outcome between the treatment groups, outcome will be defined as a three-level ordinal variable: no seizures, a single brief (<15 minute) seizure, and multiple or prolonged (>15 minutes) seizures. The analysis of this outcome variable will involve fitting an ordinal logistic regression model (proportional odds model)(78) with treatment group as the factor of interest, site and disease severity as stratification factors, and history of prior seizures and pre-existing developmental delay as covariates. The estimated treatment group odds ratio, along with its associated 95% confidence interval and p-value, will be derived from this model. If the proportional odds assumption appears to be seriously violated, a multinomial logistic regression model will be used instead (78).

Analytic Plan for Hypothesis 2:

*We hypothesize that fever treatment will reduce sequestration resulting in lower HRP2 levels based upon area under the curve during the 72-hour treatment period and an associated transient paradoxical increase in peripheral parasitemia with a higher proportion of older parasites seen*.

AUC has been selected as the outcome due to substantial between-subject heterogeneity of the level-time curves and because subjects will be at different time points in their illness in terms of treatment (some having received antimalarials as an outpatient prior to admission). As such, if a more rapid reduction in HRP2 occurs in general, it should be captured by this AUC. The analyses of the area under the HRP2 level-time curve (log-transformed) will involve fitting an analysis of covariance model with treatment group as the factor of interest, site and disease severity as stratification factors, and time since the initiation of antimalarial treatment and log-transformed HRP2 level at t=0 as covariates. The estimated treatment effect (difference in adjusted group means between the aggressive antipyretic therapy and usual care groups) and its associated 95% confidence interval, as well as a t-test for significance of the treatment effect, will be derived from this model. For subjects who do not complete the 72-hour follow-up period, the area under the HRP2 level-time curve (log-transformed) observed during the subject’s abbreviated follow-up will be multiplied by the inverse of the percentage of time that the subject was followed for the primary analysis. A secondary analysis of this outcome will be performed weighting each subject’s contribution by the time that they contribute HRP2 data.

We expect aggressive fever reduction to facilitate release of mature sequestered parasites (evident by the hemozoin they contain) from the CNS. In an exploratory analysis, the mean percentage of mature parasites across all positive blood smears for each child taken Q6 hourly will be compared in the ‘usual care’ vs. ‘aggressive antipyretic treatment’ groups using a t-test. In ‘usual care’, less than 5% of parasites typically have evidence of hemozoin deposition but the natural history of parasite hemozoin clearance is unknown.

### Interim analyses

Interim analyses for efficacy will be performed at the end of the second and third recruiting malaria seasons. An upper boundary will be determined using the O’Brien-Fleming α-spending function to control the overall probability of a Type I error at 2.5% (since only an upper boundary will be considered for early stopping).

### Methods for additional analyses (e.g. subgroup analyses)

To evaluate the effects in the study population most analogous to the BMPES population, a secondary analysis will include terms added to the above analysis of covariance model representing the main effect of subgroup (CM with CNS parasite sequestration vs. all others) and the interaction between treatment and subgroup. For purposes of this analysis, CM will be defined as having a BCS ≤2 or GCS ≤10 and CNS parasite sequestration will be defined as having an initial HRP2 ≥1700 ng/dl. This model will permit estimation of the treatment effects within each subgroup (with associated 95% confidence intervals and p-values) and a test for significance of the treatment effects between the subgroups. Exploratory analyses will be performed using the above methods for the log-transformed fever exposure outcomes. For subjects who do not complete the 72-hour follow-up period, the AUC falling above the fever cut-off during the subject’s abbreviated follow-up will be multiplied by the inverse of the percentage of time that the subject was followed for the primary analysis. Secondary analyses of these outcomes will be performed weighting each subject’s contribution by the time that they are followed, as described above for Tmax.

### Methods in analysis to handle protocol non-adherence and any statistical methods to handle missing data

For subjects who do not complete the 72-hour follow-up period, the maximum temperature observed during their abbreviated follow-up will be used for the primary analysis. A secondary analysis will be performed that is the same as the primary analysis except that the contribution of each subject’s data to the analysis will be weighted by the time that they are followed (percentage of the 72-hour follow-up period)

Plans to give access to the full protocol, participant level-data and statistical code

No later than three years from the close of the study we will deliver a completely de-identified data set to an appropriate data archive for sharing purposes.

## Oversight and monitoring

### Composition of the coordinating centre and trial steering committee

Each study site will be led by a local PI who is a senior clinician with substantial experience in pediatric malaria care. The PI will be supported by a local study coordinator. The study PI will spend some time at each study site providing logistical and clinical support for the study and coverage for the local PI as needed. A Principal Study Coordinator will provide overall management of the regulatory documents which will be maintained electronically via DropBox to assure all sites always have the most up-to-date documents. The PI will have weekly meetings with the local PIs and the Principal Study Coordinator. Larger team meetings will be held as needed.

### Composition of the data monitoring committee, its role and reporting structure

#### Data Monitoring

This study will be overseen by Local Study Monitors (LSM) at each study site and a Data and Safety Monitoring Board (DSMB). The LSMs will review adverse events and outcomes weekly. For Serious Adverse Events (SAEs, defined below) and any AEs of grade 3 or higher, the PI will be alerted immediately and the LSMs will provide expedited 48-hour review which will be circulated to the DSMB and PI on completion. LSMs are individuals with substantial experience caring for children with severe malaria. Plans for detailed management for adverse events are outlined in Table 3. Weekly LSM reports will be provided to the DSMB at ~6 monthly intervals including those provided for the annual meeting and interim analysis. If the LSM develops concerns regarding the safe continuation of the study, he/she will inform the PI to suspend further study enrollment pending DSMB review and discussion. If the DSMB identifies a concern that may require halting of enrollment or modification of the protocol, they will contact the PI and have a meeting or teleconference with other members of the study team. NINDS will be involved as appropriate, in order to come to a final decision regarding course of action.

##### SAEs include

1. Death
2. Neurologic sequelae evident at discharge
3. Aspiration requiring discontinuation of the study drug
4. The development of clinically evident bleeding or platelet dysfunction (not occult blood in urine or stool)
5. Renal injury requiring management other than simple discontinuation of the intervention
6. The development of clinically evident jaundice or an increase in bilirubin of > 5 mg/dL from baseline
7. Prolonged hospitalization potentially related to the study intervention

If study enrollment is suspended for safety reasons, the PI will notify the NIH, the supervising IRBs (the University of Rochester’s Research Subjects Review Board, The University of Malawi’s College of Medicine Research Ethics Committee, the University of Zambia’s Biomedical Research Ethics Committee) and the appropriate regulatory authorities including Malawi’s Pharmacy, Medicines and Poisons Board and Zambia’s Medicines Regulatory Authority. The DSMB consists of four clinician scientists with expertise in severe malaria in children who have worked in other African countries, plus a US-based biostatistician who has held this role on numerous DSMBs for other clinical trials. SAEs are reported to the DSMB within 48 hours, all other AEs reported to the DSMB Q 6-monthly and an interim safety analysis will be completed annually. As noted in Section 20 above, interim analyses for efficacy will also be conducted at the completion of Years 2 and 3 using an upper boundary determined by an O’Brien-Fleming α-spending function to control the overall probability of a Type I error at 2.5% (since only an upper boundary will be considered for early stopping). Interim analyses will also be performed for excess mortality in the aggressive antipyretic treatment group at the end of each of the first three recruiting malaria seasons. A lower boundary will again be determined using an O’Brien-Fleming α-spending function to control the overall probability of a Type I error. The DSMB will also carefully consider other adverse events in terms of treatment group imbalances, with particular attention paid to vomiting, recurrent hypoglycemia, elevated creatinine or bilirubin, blood transfusion requirements, identification of occult blood, and clinically evident thrombocytopenia or bleeding. Continuous variables that will be monitored include hours with lactic acidosis, maximum lactate, bilirubin, and creatinine levels, and parasite clearance time. Fisher’s exact tests (or chi-square tests) will be used to compare the treatment groups with respect to dichotomous outcomes, and t-tests (or Wilcoxon rank sum tests) will be used to compare the treatment groups with respect to continuous outcomes. A log-rank test will be used to compare the treatment groups with respect to parasite clearance time. If potential safety concerns are identified, the DSMB may consider recommending modification (or halting) of the trial. The DSMB will provide an annual review of the interim analysis through a combination of virtual and face-to-face meetings and provide a written summary report within 1 week of the annual review meeting.

An initial meeting between the study leadership and the DSMB will take place prior to enrollment of the first study subject, to review study procedures and come to agreement on the DSMB Charter. The Charter will document the role and function of the DSMB including the frequency and format of DSMB meetings, the format of reports to be prepared for DSMB review, guidelines for interim analyses for safety and efficacy, extent of blinding of the DSMB, and rules for communication among the study leadership and the DSMB.

The Department of Biostatistics and Computational Biology (DBCB) at the University of Rochester will prepare formal reports for each interim analysis for the DSMB. These reports will include appropriate introductory material (study synopsis, current status of the trial, etc.), information on recruitment and retention, subject characteristics, subject disposition, treatment discontinuations, adverse events (including serious adverse events), laboratory test results, and other safety data. This information will be provided by treatment group, where applicable.

Approximately one month before a scheduled DSMB meeting, the database will be transferred from CHeT to the DBCB using a long-established protocol, and a numbered version of the analytic database will be created by the DBCB. All DSMB reports will refer to the specific version number of the database that is used in their preparation, as well as the date on which the database was created. The designated DBCB programmer who generated the randomization plan for the trial will prepare the DSMB reports. The treatment groups in the report will be coded as “A” and “B”, which can be linked to the identities of the actual treatment groups if requested by the DSMB.

### Adverse event reporting and harms

#### Harms

CM is associated with high rates of morbidity and mortality. The expected outcomes related to the disease itself include severe anemia requiring blood transfusions, hyperlactatemia, acidosis, prolonged coma, hypoglycemia, severe thrombocytopenia, hyponatremia, extreme hyperpyrexia, seizures, and, neurologic sequelae including blindness, hemiparesis, hypotonia, and mutism. Death is also a common outcome, occurring in 16% of pediatric CM patients on the QECH Research Ward over the past 5 years. All study subjects will receive treatment and management for these conditions consistent with the WHO’s guidelines for the management of severe malaria.

Known potential harms from the Intervention are listed below

a. Aspiration: Study subjects will be closely monitored clinically for signs of aspiration including Q4-hourly assessments of oxygen saturation. The Q4-hourly assessments are intended to be done only if coma score is sub-normal, otherwise this metric need be done only per hospital ward protocol. If oxygen saturations are below 95%, the ward attending will be notified to make a physician-level assessment (auscultation, urgent PCV, etc.) and determine if aspiration is the potential underlying cause. If oxygen saturation declines and remains lower than the pre-aspiration baseline, the NGT will be placed to suction to remove any remaining gastric contents, the intervention will be discontinued and the NGT removed. Otherwise, the decision to discontinue the study drug intervention is at the discretion of the attending physician.
b. Complications of NGT placement and maintenance: If placement/maintenance of the NGT by experienced staff is difficult and results in nasal trauma and/or significant epistaxis and the study subject is comatose and/or unable to swallow, the intervention will be discontinued and the NGT removed. Necessary measures to ameliorate any bleeding will be taken.
c. Vomiting: Although common in the early stages of malaria, once comatose study participants with CM do not usually vomit. If vomiting occurs in a comatose study participant, the NGT will be placed to suction to remove the remaining gastric contents, oxygen saturations will be assessed, and once gastric contents have been removed the NGT will be removed and the intervention will be discontinued.
d. Acidosis: Lactic acidosis is common on admission but generally resolves within 24 hours.
e. Hyperbilirubinemia: Liver dysfunction is common in CNS malaria. Children with a bilirubin on admission of above 3 or those with frank jaundice are excluded. The study intervention will be halted if after enrollment the bilirubin escalates >5mg/dL or the child develops clinical jaundice.
f. Renal insufficiency: If the creatinine increased by >0.5 mg/dL from baseline or a doubling of the baseline creatinine, the study intervention will be stopped.
g. Increased risk of bleeding: Thrombocytopenia is common during malaria infections. The use of ibuprofen may decrease platelet function and could therefore increase the risk of abnormal bleeding. Children with any history of thrombocytopenia prior to the acute malaria illness will be excluded. Structured clinical screening will also be conducted to assure anyone with evidence of abnormal bleeding is excluded from enrolment. During the study, children will be evaluated twice daily by an experienced clinician who will repeat the structured assessments to identify any clinical evidence of abnormal bleeding/poor clotting and the intervention will be halted in any child that develops signs/symptoms.

### Auditing

Prior to the initiation of the study, harmonization training sessions will be conducted for clinician co-investigators, the investigators, and their study coordinators for each site. This meeting will include a detailed discussion of the protocol, performance of study procedures, CRF completion, simulation of study procedures and specimen collection methods, as applicable. Subsequent detailed local training sessions with ward nurses and clinicians will be held at each site.

Prior to commencing enrolment, the University of Malawi Research Support Centre (RSC) will conduct an external audit of the study in Malawi and a certified RSC auditor will conduct a similar audit of the Zambian site to assure the clinical trial is in compliance with Good Clinical Practices.

After completion of the data entry process, computer logic checks will be run to check for such items as inconsistent study dates and outlying laboratory values. Any necessary correction will be made to the database and documented via addenda or audit trail. A manual review of selected line listings will also be performed at the end of the study.

The Principal Study Coordinator will assist the PI in assuring protocol compliance, ethical standards, regulatory compliance and data quality at the clinical sites. The coordinator will provide real time daily review of all study files to assure protocol compliance and documentation completeness. The study sites may also be subject to quality assurance audits by the NINDS or its designees and appropriate regulatory agencies. Audits may include reviews of paper or electronic records.

### Research ethics approval

In addition to the oversight provided at each site by the local study monitors, the study has been approved by the following Institutional Review Boards:

- The University of Zambia’s Biomedical Research Ethics Committee FWA00000338 (UNZA BREC)-initially approved on 17 August 2018, renewed on 16 November 2021, approval number 003-06-18.
- The University of Malawi’s College of Medicine Research Ethics Committee FWA00011868 (COMREC)-initially approved on 08 November 2017, renewed on 09 November 2021, approval number P.10/17/2298
- The University of Rochester’s Research Subjects Review Board FWA00009386 (RSRB) - initially approved 27 April 2017, renewed on 16 November 2021, approval number RSRB 000667717

An intra-institutional agreement was generated between the University of Rochester’s RSRB and Michigan State University’s (MSU) Biomedical Institutional Review Board FWA00004556 (BIRB). Michigan State University’s BIRB and Boston Children’s Hospital (BCH) IRB have agreed to rely on University of Rochester’s RSRB.

While in Malawi, the MSU PI is responsible for overseeing subject recruitment and data collection at Queen Elizabeth Central Hospital. As part of data cleaning process, he will also have access to data with patient identifiers.

While in Zambia, the BCH PI will serve as a clinician. Her responsibilities also include data collection and conducting EEG’s on study patients. As part of data cleaning, she will also have access to data with patient identifiers.

Our office takes responsibility for the required IRB reports (the University of Rochester’s Research Subjects Review Board, The University of Malawi’s College of Medicine Research Ethics Committee, and the University of Zambia’s Biomedical Research Ethics Committee) within Malawi and Zambia. Regular communications between the MSU PI and BCH PI, Local Safety Monitoring (LSM) and the Lead PI ensure that data collected and contact information is up-to-date.

No human subjects contact will occur at any of the US-based sites. All data exported to the US is de-identified prior to transfer. All RSRB determinations regarding this work will be shared with the non-affiliated researchers in a timely fashion.

All laboratory specimens, evaluation forms, reports, and other records that leave the participants enrollment sites will be identified only by the study ID number to maintain subject confidentiality. All paper records will be kept in a secure study office, within locked file cabinet. All computer entry and networking programs will be done using IDs only. Clinical information will not be released without written permission of the subject or the subject’s parent or guardian, except as necessary for monitoring by the supervising IRBs (the University of Rochester’s Research Subjects Review Board, The University of Malawi’s College of Medicine Research Ethics Committee, the University of Zambia’s Biomedical Research Ethics Committee) and regulatory agencies, (Medicines and Poisons Board in Malawi and Zambia Medicines Regulatory Authority), the sponsor, and/or the sponsor’s designee. Any presentation, abstract, or manuscript will be made available to the sponsor and IRBs as part of routine annual reports.

### Protocol amendments

The protocol included in this submission, which was developed in accordance with the 2013 SPIRIT Guidelines, and any subsequent modifications will be reviewed and approved by the ethical review boards listed in Section 24, the local study monitors, and an Advisory Committee which will include leadership from the Department of Pediatrics in both African institutions. Any modifications to the protocol which may impact on the conduct of the study, potential benefit of the patient or may affect patient safety, including changes of study objectives, study design, patient population, sample sizes, study procedures, or significant administrative aspects will require a formal amendment to the protocol. Such an amendment will be agreed upon by the University of Rochester, NINDS, and the Advisory Committee, and approved by the IRB prior to implementation, and notified to the health authorities in accordance with local regulations. Administrative changes of the protocol are minor corrections and/or clarifications that have no effect on the way the study is to be conducted. These administrative changes will be agreed upon by the Advisory Committee and will be documented in a memorandum. The IRB may be notified of administrative changes at the discretion of the PI

## Dissemination plans

The primary outcome of this study will be submitted for publication no later than one year after study completion. This manuscript, as well as any other manuscripts, posters, or abstracts, must be approved by the publication committee for approval prior to submission. Financial support from the NINDS will be acknowledged in all publications

## Discussion

Despite substantial evidence that hyperthermia in the setting of acute CNS injury generally worsens neurologic damage as well as observational data in pediatric CNS malaria showing the same association, malaria care guidelines today recommend a single antipyretic agent and treatment initiation only after a significant fever (T ≥ 38.5°C) occurs. The clinical trial proposed here seeks to challenge this practice paradigm by evaluating the fever-reduction efficacy of more aggressive antipyretic use while also taking advantage of a relatively new method for quantifying total parasite burden (HRP2) to further characterize malaria severity and elucidate the impact of antipyretics on parasite sequestration and clearance. This study will also assess the efficacy of aggressive antipyretic use in deterring acute symptomatic seizures during the malaria infection. The trial is novel in the provision of prophylactic treatment, the proposed use of combining two agents, and the select patient population—namely children with CNS malaria rather than uncomplicated malaria as were included in prior antipyretic malaria trials.

## Trial status

Protocol version # 5.3, issue date: 21 Oct 2020

Authors: Karl B. Seydel, Gretchen L. Birbeck, Malcolm Molyneux

Revision Chronology: GLB revision to version 3.0 30Sep2016

The trial is currently recruiting and started on 7Jan2019. We anticipate recruitment will be completed by July 2022.

## Data Availability

No datasets were generated or analysed during the current study. All relevant data from this study will be made available upon study completion

## Abbreviations

PI: Principal Investigator
CM: Cerebral Malaria
M/SM: Malaria and Seizures
HRP2: Histidine Rich Protein
qPC: quantitative Parasite Count
AUC: Area under Curve
PRW: Pediatric Research Ward at Queen Elizabeth Hospital in Blantyre
PICU: Pediatric Intensive Care Ward (PICU) at University Teaching Hospital in Lusaka, Zambia
UR: University of Rochester
BCH: Boston Children’s Hospital
MSU: Michigan State University
LSM: Local Study Monitor
DSMB: Data and Safety Monitoring Board
SAEs: Serious Adverse Events
UNZA BREC: The University of Zambia’s Biomedical Research Ethics Committee
COMREC: The University of Malawi’s College of Medicine Research Ethics Committee
RSRB: The University of Rochester’s Research Subjects Review Board

## Declarations

None of the investigators on the trial declare any competing interests.

### Authors’ contributions

GB conceived of the study. GB and KBS initiated the study design, Moses C will help with study implementation. Michael M will complete the initial statistical analysis. All authors contributed to refinement of the study protocol.

### Availability of data and materials

The DBCB will oversee the intra-study data sharing process, with input from the Principal Investigator.

All Investigators (both US and host country) will be given access to the cleaned data sets. Individual country investigators will have direct access to their own site’s data sets and will have access to other site’s data by request. To ensure confidentiality, data dispersed to project team members will be blinded of any identifying participant information

### Ethics approval and consent to participate

Written informed consent will be obtained from all participant’s parents or guardians. Since participants are by definition neurologically impaired during the 72-hour study period, assent will not be sought.

Study approval will be obtained from the following oversight entities prior to the conduct of this work:

- The University of Zambia’s Biomedical Research Ethics Committee FWA00000338 (UNZA BREC)
- The University of Malawi’s College of Medicine Research Ethics Committee FWA00011868 (COMREC)
- The University of Rochester’s Research Subjects Review Board FWA00009386 (RSRB)
- An intra-institutional reliance agreement will be generated between the University of Rochester’s RSRB and Michigan State University’s (MSU) Biomedical Institutional Review Board FWA00004556 (BIRB).
- The Zambian Medicines Regulatory Authority (ZAMRA)
- The Pharmacy and Medicines Regulatory Authority (PMRA) in Malawi
- The National Health Research Authority (NHRA) in Zambia

### Consent for publication

Consent forms in English, Nyanja and Chewa as well as information sheets provided to parents/guardians are available

